# Probiotic responder identification in cross-over trials for constipation using a Bayesian statistical model considering lags between intake and effect periods

**DOI:** 10.1101/2022.03.14.22272054

**Authors:** Shion Hosoda, Yuichiro Nishimoto, Yohsuke Yamauchi, Takuji Yamada, Michiaki Hamada

## Abstract

Recent advances in microbiome research have led to the further development of microbial interventions, such as probiotics and prebiotics, which are potential treatments for constipation. However, the effects of probiotics vary from person to person; therefore, the effectiveness of probiotics needs to be verified for each individual. Individuals showing significant effects of the target probiotic are called responders. A statistical model for the evaluation of responders was proposed in a previous study. However, the previous model does not consider the lag between intake and effect periods of the probiotic. It is expected that the lag exists when probiotics are administered and when they are effective. In this study, we propose a Bayesian statistical model to estimate the probability that a subject is a responder, by considering the lag between intake and effect periods. In synthetic dataset experiments, the proposed model was found to outperform the base model, which did not factor in the lag. Further, we found that the proposed model could distinguish responders showing large uncertainty in terms of the lag between intake and effect periods.

## 1 Introduction

Recent advances in microbiome research have resulted in the rapid development of microbial interventions, such as probiotics and prebiotics, which are potential treatments for constipation [1]. Probiotics are living microbes that benefit the host when ingested in sufficient quantities and are reported to improve defecation frequencies and treat constipation [1, 2]. The effects of probiotics vary from person to person [3]. Individuals exhibiting significant effects of probiotics are called “responders” [4]; each responder exhibits a significant effect of a different probiotic. That is, different responders respond to different probiotics, and individual differences make it difficult to evaluate the effects of probiotics. Therefore, experimental designs used for probiotic research should consider the individual differences between subjects.

One type of sophisticated experimental design is a cross-over trial, in which each subject takes both the target probiotic and a placebo. A cross-over trial comprises the following steps: (1) Each individual is first administered a capsule containing the target probiotic or placebo for several days. (2) After a washout period, which lasts several weeks and is set to remove the effects of the former capsule, each individual is administered another capsule (containing either a probiotic or a placebo) for a specific period. Cross-over trials are widely applied in various fields of research, including research related to probiotics [5], prebiotics [6], neurorehabilitation [7], and spinal manipulation [8]. The main advantage of cross-over trials is that they enable the evaluation of individual differences [9], which are supposed to be determined by attributes such as gender, genetics, and habits. Accordingly, datasets obtained from a cross-over trial require a reasonable analysis method that considers individual differences.

An approach to estimate individual differences has already been conducted in a previous study. Nakamura *et al*. evaluated improvements in defecation frequencies using a Weibull regression model [10]. They revealed individual differences in the improvement of defecation frequency by grouping subjects into three groups: strong responders, weak responders, and non-responders. However, their model used the unreasonable assumption that the effects of the target probiotics start on the day the probiotic is administered to the subject. A previous study suggested that orally ingested material should be excreted for one or more days [11]. In addition, it has been estimated that it can take more than ten hours for microbes to increase dramatically [12] and at least two days for food to alter the gut microbiome [13]. Therefore, analyses that do not consider the lag between intake and effect periods can lead to a misidentification of responders, especially in short-term intervention experiments.

In this study, we propose a Bayesian statistical model for estimating the efficacy of a target probiotic in improving defecation frequency, considering the lag between intake and effect periods (Figure 1) and individual differences in a cross-over trial dataset. This study aimed to identify responders accurately using cross-over trial datasets where the lag between intake and effect periods exists. The proposed model is based on the segmented linear regression model, which represents each periodic term using linear regression, and has discrete parameters for lag days. The proposed model evaluates the cumulative sum of the number of times a subject defecates. An individual can be evaluated based on the posterior probability that the individual is a responder to probiotics. With the proposed model, we estimated whether each subject is a responder using synthetic datasets and the real dataset used in the previous study [10]. We compared the results of the proposed model with those of a base model that did not consider the lag period. Our analysis showed that considering the effect of time lag was useful in the synthetic dataset experiments. Real data experiments showed that the proposed model estimated the posterior distribution considering the effect lag and led to different conclusions from those of the base model. We found that the proposed model could eliminate uncertain responders (responders whose response to a probiotic is uncertain) according to the lag between intake and effect periods.

**Figure 1:**
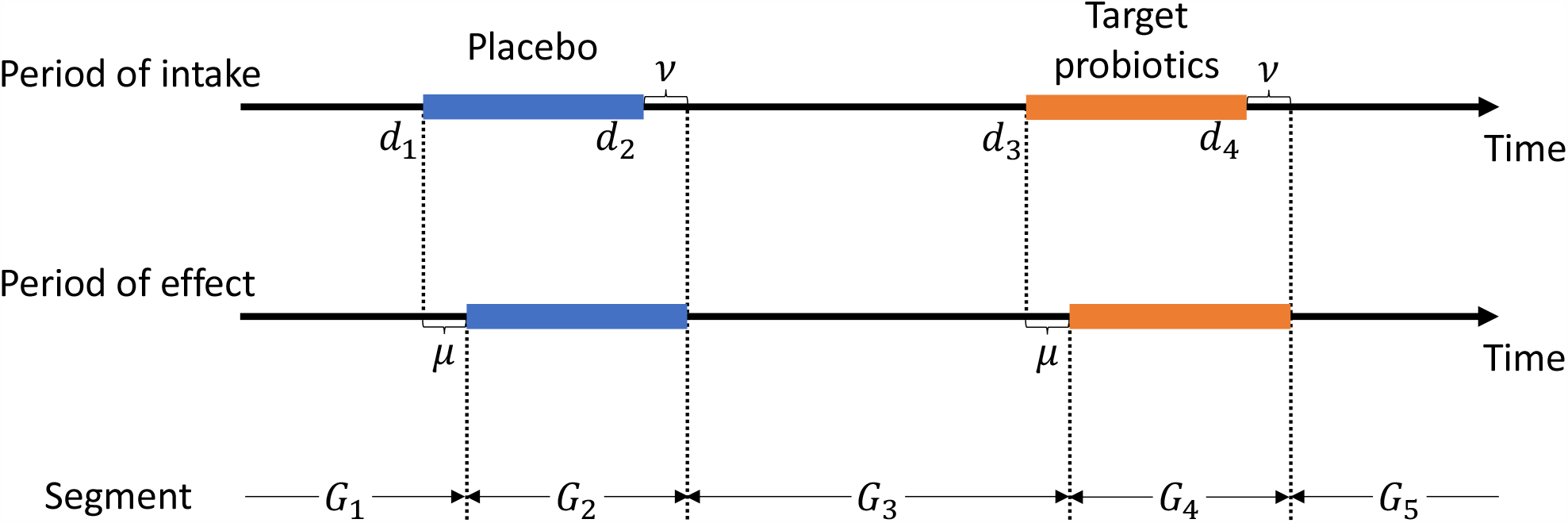
Schematic illustration of the effect of lag in cross-over trials. Blue and orange boxes represent the placebo and target probiotics periods, respectively. *d*_1_, *d*_2_, *d*_3_, and *d*_4_ represent the start day index of the first capsule, the end day index of the first capsule, the start day index of the second capsule, and the end day index of the second capsule, respectively. *μ* and *ν* are the lag of effect start and end, respectively. *G*_*t*_ shows the term of the day in the *t*-th segment. The time when an effect of the probiotic was observed was delayed compared to the time the probiotic was ingested. In this case, the subject was first administered the placebo capsule and then the target probiotic capsule.

## 2 Materials and methods

### 2.1 Overview

Here, we provide an overview of the proposed model. The proposed model requires a dataset represeting the cumulative sum of the number of defecation events collected from a cross-over trial. Figure 1 shows the schematic illustration of the respective cross-over trial whose dataset was used in this study. The terms of the trial are divided into several periodic terms with respect to capsule intake/effect and are denoted as “segments.” Here, *S* = 5 is used in this figure and the dataset used in this study, where *S* is the number of segments. The proposed Bayesian statistical model is based on segmented linear models for the cumulative sum of the number of defecation events. The proposed model can consider the lag between intake and effect periods (*cf*. Section 1) using the number of lag days as discrete parameters.

### 2.2 Generative process

Here, we describe the generative process of the proposed model. The proposed model demands the cumulative sum of the defecation frequency *y*_*i*_(*i* = 1 … *N*), where *N* is the number of days in the entire trial term. This model is for one subject. Let *α* ∈ ℝ, *η* ∈ ℝ, and 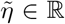 be the logarithmic defecation frequency during normal periods, the effect of target probiotics, and the effect of capsules, respectively. Here, the effect of the capsule indicates the effects of ingesting the capsule, regardless of its content. That is, the effect of the capsule is observed in both the target probiotic and placebo periods. Although 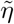 is utilized to denote the effect of “the capsule,” the proposed model does not rely on the form of the target probiotics or the placebo. The prior distributions of *α, η*, and 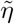 are as follows.

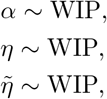

where WIP denotes the weakly informative prior distribution. We use Cauchy(0, 10) as the weakly informative prior distribution in this study, where Cauchy(*x*_0_, *γ*) denotes the Cauchy distribution with the location parameter *x*_0_ and scale parameter *γ*. Let *μ* and *ν* be the lag of effect start and end, respectively. *μ* and *ν* are shared by the effects of capsules and probiotics. Specifically, the effects of the capsules and probiotics emerged *μ* days after the subject ingested the capsule and expired *ν* days after the subject stopped taking the capsule. Here, we assume that *μ* and *ν* are up to several days long using the following prior distributions:

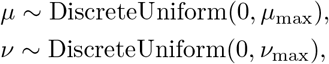

where DiscreteUniform(*a, b*) denotes the discrete uniform distribution with minimum value *a* and maximum value *b*, and *μ*_max_ and *ν*_max_ are the maximum values of *μ* and *ν*, respectively. We used *μ*_max_ = *ν*_max_ = 5 in this study. We modified the segment in which the subject ingested the capsule using *μ* and *ν* as follows:

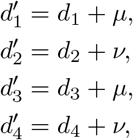

where *d*_1_, *d*_2_, *d*_3_, and *d*_4_ represent the start day index of the first capsule, the end day index of the first capsule, the start day index of the second capsule, and the end day index of the second capsule, respectively; and 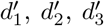, and 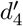 indicate the effect start day index of the first capsule, the effect end day index of the first capsule, the effect start day index of the second capsule, and the effect end day index of the second capsule, respectively. Let *O, P*, and *T* be the sets of day indices in the normal, placebo, and target probiotic periods, respectively. *O, P*, and *T* are given by

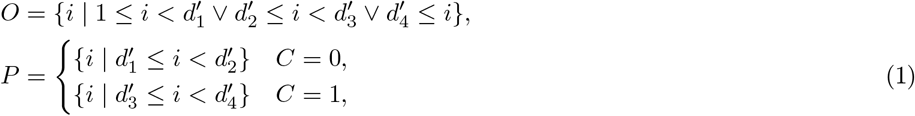

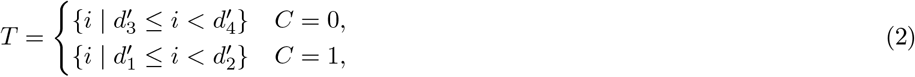

where *C* indicates the cross-over type, which shows the order of the target probiotic and placebo capsules. The subject was first administered the placebo and then the target probiotic when *C* = 0, and the subject was first administered the target probiotic and then the placebo when *C* = 1. Let *β*_*i*_ be the rate of increase in the cumulative sum of the defecation frequency on the *i*-th day. *β*_*i*_ depends on the period in which the *i*-th day lies, as given below:

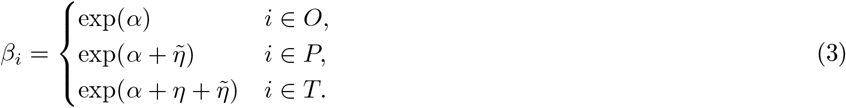

The intercept of the *i*-th segment *γ*_*i*_ is defined as

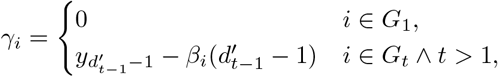

where *G*_*t*_ is the set of the day indices in the *t*-th segment. Here, *γ*_*i*_ is calculated such that the regression line passes through the observation point 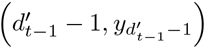. This calculation enables the precise evaluation of the increase in the cumulative sum of the defecation frequencies in each segment. *γ*_*i*_ in the first segment equals zero because the cumulative sum of defecation frequencies is 0 before the first day. Here, we considered the placebo effect of defecation frequencies because placebo effects have been confirmed in previous studies [14, 15]. The distribution of the *i*-th day cumulative sum of defecation frequencies *y*_*i*_ is as follows:

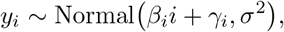

where

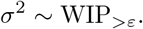

Here, WIP_*>ε*_ denotes the truncated weakly informative prior distribution, whose domain of definition is *x > ε* with a random variable *x*. We used 0.1 as *ε*.

### 2.3 Parameter estimation

We estimated the posterior distribution of the parameters of the proposed model using the No-U-Turn-Sampler (NUTS) [16], which is a Markov chain Monte Carlo (MCMC) method. Because the NUTS can sample only continuous parameters, we estimated the following posterior distribution marginalized with respect to *μ* and *ν*:

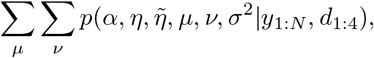

where *·*_1:*N*_ denotes the set 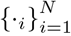. We implemented a parameter estimation algorithm using PyStan (https://github.com/stan-dev/pystan). We used five chains of MCMC and then sampled parameters 1000 times randomly for each chain and discarded the first half of the samples as burn-in samples, which were supposed to depend on the initial sample. We used 15 as the maximum tree depth in the NUTS algorithm (called the “max_treedepth” option in the PyStan library). The other hyperparameters were set by default.

### 2.4 Evaluation by scoring improvement of defecation frequency

We defined the following defecation frequency improvement (DFI) score DFI(*μ, ν*):

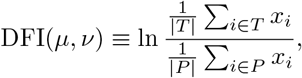

where *x*_*i*_ is the defecation frequency of the *i*-th day, and *T* and *P* are defined by Eq. (1) and Eq. (2), respectively, for the score parameters *μ* and *ν*. The DFI score indicates the log ratio of the defecation frequency in the target probiotic period to that in the placebo period.

### 2.5 Synthetic data experiment

We generated synthetic datasets and estimated the parameters using these datasets to evaluate the performance of the proposed model. We generated *α, η*, 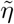, *μ*, and *ν* using the following distribution:

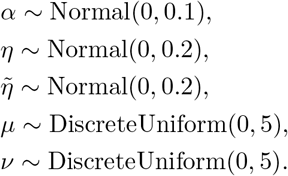

After computing *β*_*i*_ using *α, η*, 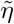, *μ*, and *ν* (*cf*. Eq. (3)), the number of days between the *l*-th and *l* + 1-th defecation events for subject *v*_*l*_ ∈ ℝ was obtained as follows:

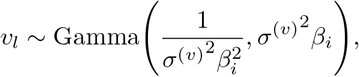

where Gamma(*a, b*) denotes the gamma distribution with the shape parameter *a* and scale parameter *b* and 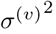 is the variance of the interval. The mean and variance of this gamma distribution were 1*/β*_*i*_ and 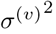, respectively. We generated *v*_l_ until ∑_*l*_ *v*_*l*_ exceeded the number of days in each segment and obtained *y*_*i*_ by transforming the defecation intervals. We randomly generated datasets 1000 times and estimated the posterior distributions of the parameters once for each dataset. We used (*d*_1_, *d*_2_, *d*_3_, *d*_4_) = (29, 43, 71, 85), (51, 76, 126, 151), (101, 151, 251, 301), *N* = 85, 151, 301, 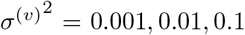, and *C* = 0 in one half of the subjects and *C* = 1 in the other half of the subjects for each dataset.

### 2.6 Real data experiment

We used a real dataset from a previous study [10], which conducted a randomized double-blind controlled cross-over trial. Twenty subjects received *Bifidobacterium longum* capsules in the experiment. Eleven subjects were administered placebo capsules from day 29 to 42 and the target probiotic capsules from day 71 to 84 (*C* = 0), and the remaining nine subjects were administered them in the reverse order (*C* = 1). That is, (*d*_1_, *d*_2_, *d*_3_, *d*_4_) = (29, 43, 71, 85) was used in the trial. Subjects reported their defecation frequencies every day in the trial term. See [10] for the details of the trial.

### 2.7 Bayesian beta regression of responder probability on the microbial relative abundances

We conducted regression analysis using the beta regression model [17]. Let *r*_*i*_ be the response probability of the *i*-th subject. The Bayesian beta regression model represents *r*_*i*_ using the standardized relative abundances of the bacteria in the *i*-th subject shortly before the start of capsule administration, which is denoted by **m**_*i*_. **m**_*i*_ is the *D*-dimensional vector, and *D* is the number of the different bacteria. The generative process is as follows:

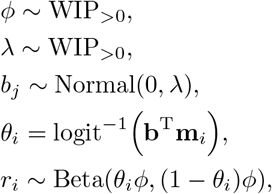

where *ϕ* is the precision parameter obtained by reparameterizing the beta distribution parameters. *λ* is the regularization parameter. **b** = (*b*_1_, …, *b*_*D*_)^T^ is the regression parameter vector; logit^−1^(*·*) is the inverse-logit function; and Beta(*a, b*) denotes the beta distribution with the shape parameters *a* and *b*. Because the domain of the definition of the beta distribution does not include zero or one, we added 10^−5^/−10^−5^ to *r*_*i*_ when *r*_*i*_ is zero or one. The same method as in Section 2.3 was used for the parameter estimation.

### 2.8 Microbiome data

The 16S rRNA sequence data were obtained from the DDBJ DRA(DRA006874). QIIME2 (version 2019.10) was used for the 16S rRNA gene analysis [18]. In the analytical pipeline, sequence data were processed using the DADA2 pipeline for quality filtering and denoising (options: –p-trunc-len-f 150 –p-trunc-len-r 190–p-max-ee-f 3.0 –p-max-ee-r 3.0) [19]. The filtered output sequences were assigned to different taxa using the “qiime feature-classifier classify-sklearn” command with the default parameters. Silva SSU Ref Nr 99 (version 132) was used as the reference database for taxonomy assignment [20]. For the regression analysis, we used only those taxa with a non-zero abundance in at least 15 subjects.

## 3 Results

### 3.1 Performance evaluation with synthetic datasets

We evaluated the performance of the proposed model under various conditions using synthetic datasets (*cf*. Section 2.5). To verify the accuracy of *η*, we compared the estimated and true values (Supplementary Figure S1). In the case of 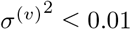, the proposed model could accurately estimate *η*. 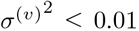, the standard deviation *σ*^(*v*)^ ≤ 0.1, means that defecation events with the one-sigma error are within ±2.4 hours (*cf*. Section 2.5).

We also verified the estimation accuracy of *μ* and *ν*. Figure 2 shows the sum of the probabilities for each true *μ* and *ν* to evaluate the uncertainty of the estimation. The diagonal elements in Figure 2def, which shows the results of *ν*, are high in the 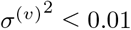 experiments. However, as shown in Figure 2abc, which shows the results for *μ*, the proposed model tends to overestimate the *μ* value.

**Figure 2:**
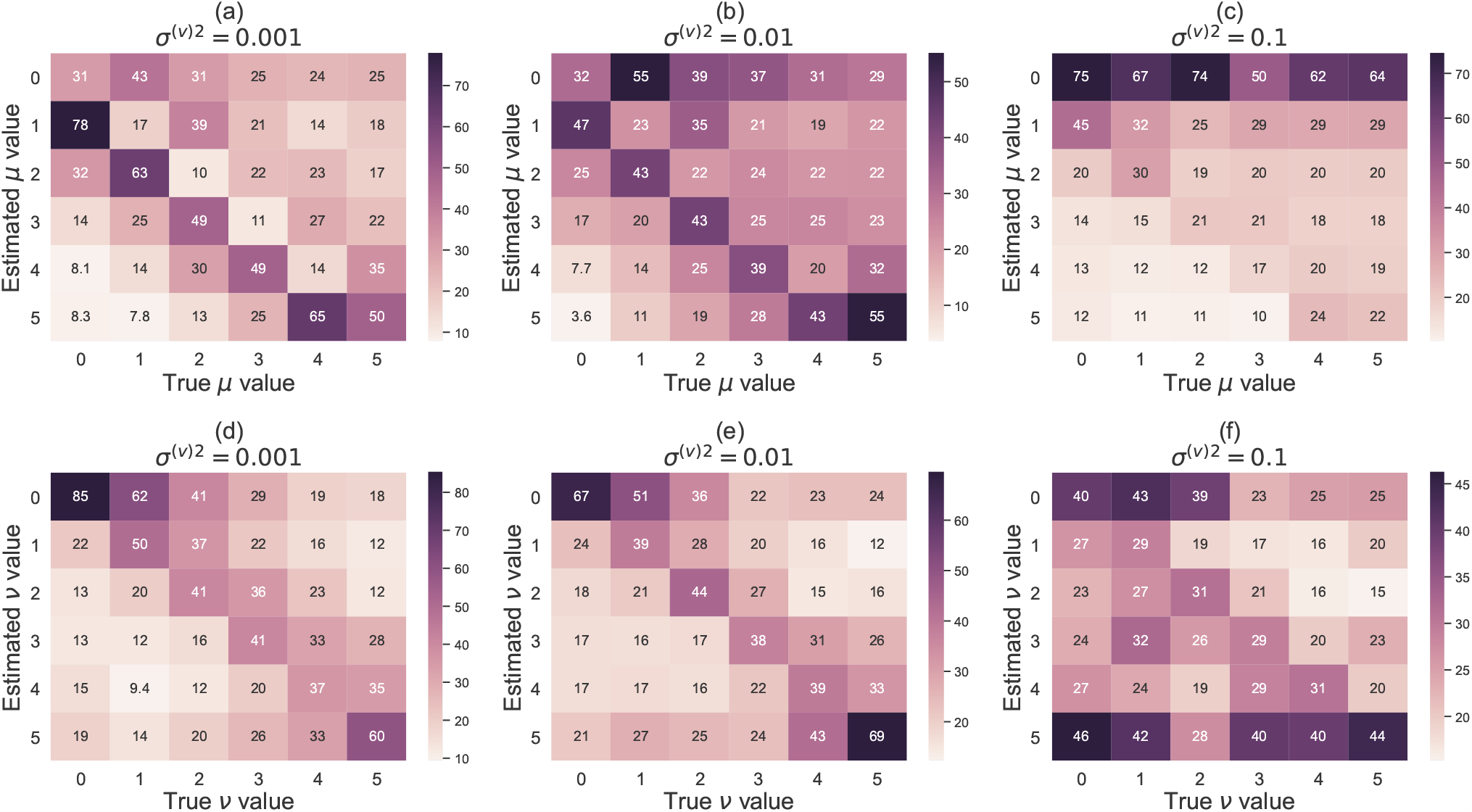
The heat map of the true *μ, ν* and *μ, ν* estimated by the proposed model for each synthetic dataset, where the number of observation points is the same as that of the real dataset. The *x*- and *y*-axes indicate the true *μ, ν* and estimated *μ, ν* values, respectively. Each column represents the sum of the probabilities of each estimate for the subject, whose true value is that in the column. a, b, and c indicate the *μ* results when 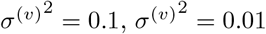, and 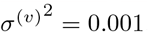, respectively. d, e, and f indicate the *ν* results when 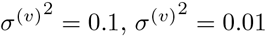, and 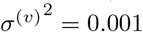, respectively.

To evaluate the performance improvement by considering the lag between intake and effect periods, we identified responders based on the posterior distributions. Here, we defined responders as the subjects with *η >* 0. Figure 3 shows the receiver operating characteristic (ROC) curve of the proposed and base models. We used the proposed model with *μ*_max_ = *ν*_max_ = 0 (*cf*. Section 2.2), which does not consider the lag, as the base model. The proposed model outperformed the base model, and the effectiveness of considering the lag was demonstrated in the case where a lag exists. Figure 3 also shows the performance for each threshold of the posterior probability of *η >* 0. In the case of 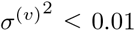, identification with a threshold of 0.95 showed a low false positive rate.

**Figure 3:**
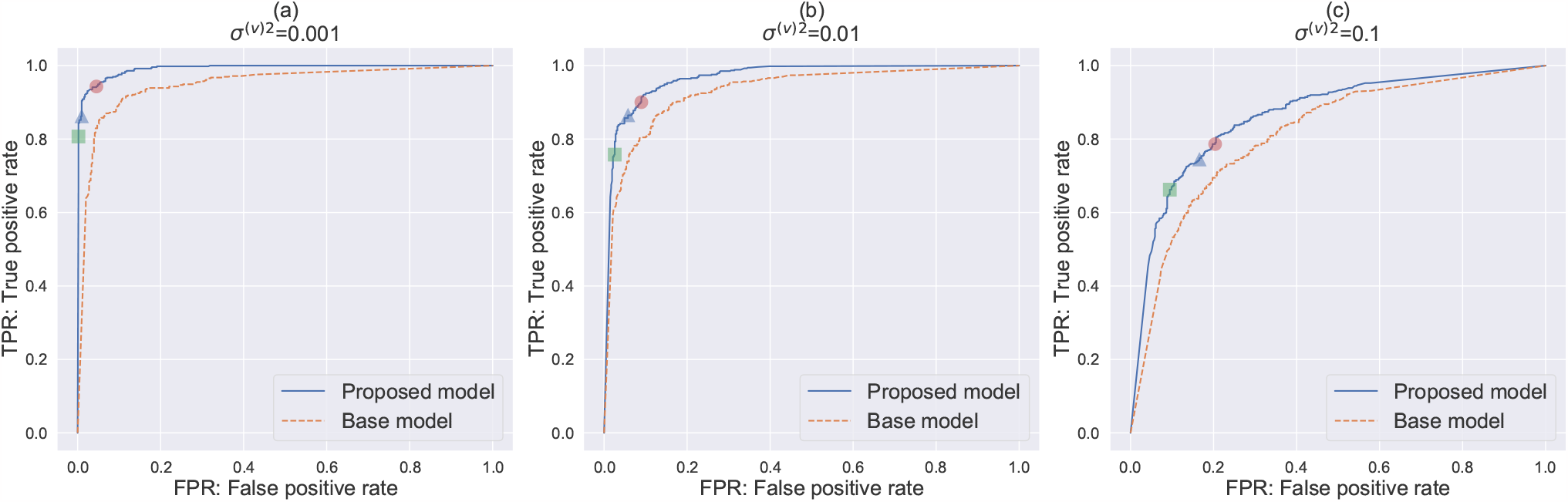
The ROC curve for identifying responders based on the estimated posterior distributions of *η* in the synthetic datasets of *N* = 85 and (*d*_1_, *d*_2_, *d*_3_, *d*_4_) = (29, 43, 71, 85) using the proposed model (*μ*_max_ = *ν*_max_ = 5) and the base model (*μ*_max_ = *ν*_max_ = 0). The *x*- and *y*-axes indicate the false positive and true positive rates, respectively. The blue and orange dashed lines indicate the results for *μ*_max_ = *ν*_max_ = 5 and *μ*_max_ = *ν*_max_ = 0, respectively. The red circle, blue triangle, and green square indicate the performance of responder identification with the threshold of the probability of *η >* 0 0.5, 0.7, and 0.95, respectively, when *μ*_max_ = *ν*_max_ = 5.

### 3.2 Responder evaluation using a real dataset

We conducted an experiment using a real dataset (*cf*. Section 2.6). To evaluate the effect of the target probiotic on each subject, we visualized the estimated posterior distribution of *η* (Figure 4). Subjects MO04, MO05, MO10, and MO16 exhibited high values of *η*, which mean high effectiveness of probiotics. In contrast, subjects MO06 and MO18 exhibited low values of *η*. The 95% Bayesian credible intervals of subjects MO02, MO04, MO05, MO06, MO08, MO10, MO12, MO13, MO16, MO18, and MO23 did not include zero. The posterior distributions of *μ* and *ν* are shown in Supplementary Figure S2. We can see that the estimated values of *μ* and *ν* vary from person to person.

**Figure 4:**
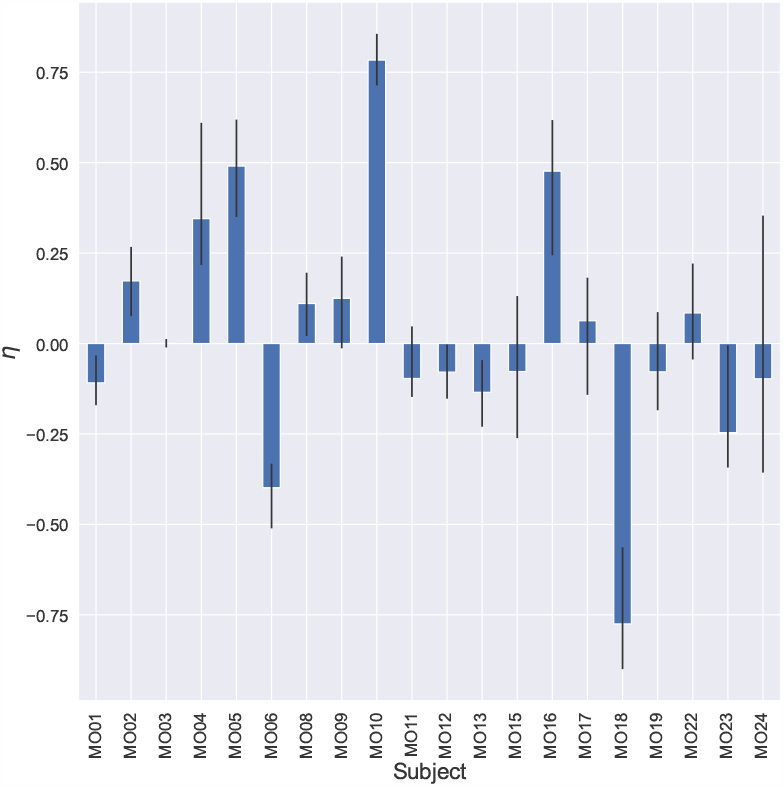
Estimated posterior distributions of *η* for each subject. The *x*- and *y*-axes indicate the subjects and *η* values, respectively. The bar shows the median of the posterior distribution. The error bars represent the 2.5% and 97.5% percentiles.

We also examined the estimated probability that each subject was a responder (Figure 5). We counted the number of samples that satisfied *η >* 0 and computed the ratio of the count to the number of all samples as the posterior probability. The probabilities of subjects MO02, MO04, MO05, MO08, MO09, MO10, and MO16 exceeded 0.95. In a previous study, subjects MO04, MO05, and MO10 were reported as responders, whereas subject MO16 was a non-responder [10]. Supplementary Figure S3 shows the posterior distributions of *η* estimated by the base model, which did not consider the lag (*μ*_max_ = *ν*_max_ = 0). The Bayesian credible interval of subject MO16 also did not include zero, but the median was estimated to be lower than that of the proposed model. The results for subjects MO22 and MO24 showed large differences between the proposed and base models. The median values of the base model were larger than those of the proposed model, and their identification of responders based on 95% Bayesian credible intervals led to different conclusions (Figure 5 and Supplementary Figure S4).

**Figure 5:**
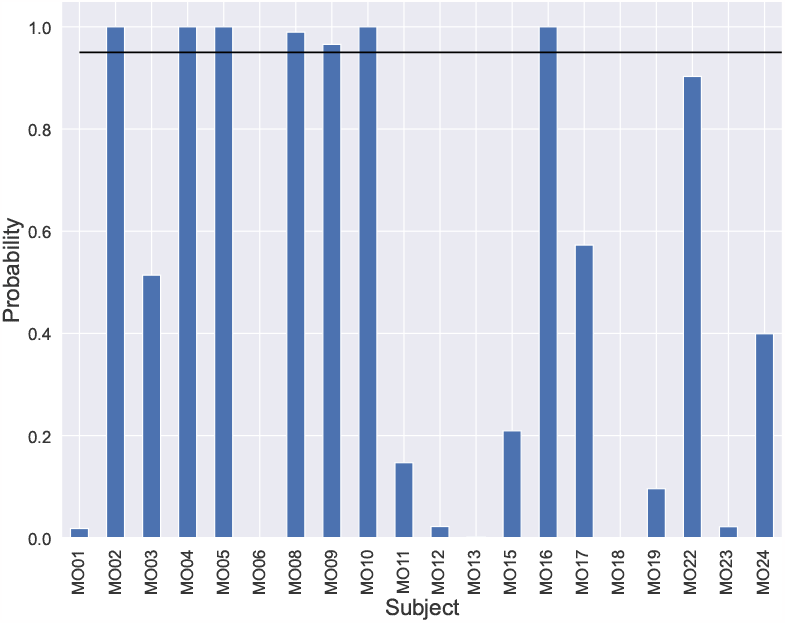
The probability that each subject is a responder based on the posterior distribution. The *x*- and *y*-axes indicate the subjects and probabilities of *η >* 0, respectively. The horizontal line indicates that the probability equals 0.95.

To verify the consistency between the posterior distributions and the used dataset, we evaluated the improvement in the defecation frequency using scoring (*cf*. Section 2.4). Figure 6 shows the DFI score of subjects where *μ* = 0 … 5 and *ν* = 0 … 5.

**Figure 6:**
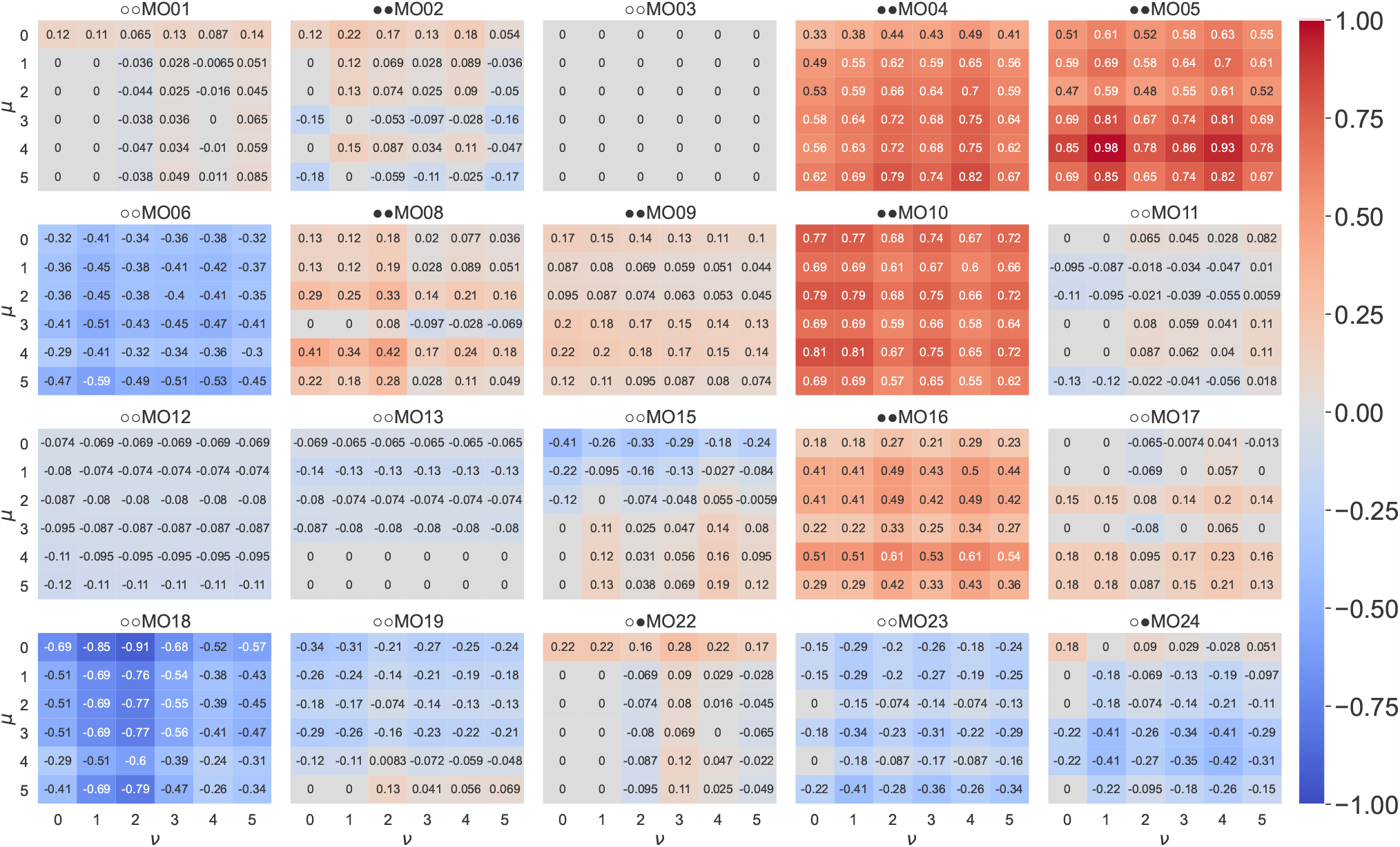
DFI scores (*cf*. Section 2.4) of all subjects. The title of each panel indicates the subject and the result of the responder identification by the proposed and base models. The left and right circles indicate the proposed and base model results, respectively. The filled circle indicates that the subject is identified as a responder, and the open circle indicates that the subject is not. The *x*- and *y*-axes indicate *ν* and *μ* of the score parameters, respectively. Each value indicates the score. A darker color indicates a higher score.

While the DFI scores of *μ* = 0 and *ν* = 0 for subject MO24, which was identified as a responder by the base model, were 0.18, the DFI scores for *μ* ≠ 0 and *ν* ≠ 0 were less than 0. Therefore, subject MO24 did not show an improvement in the defecation frequency when a lag in the effect period existed, and the proposed model reflected the specifications of subject MO24. We also examined the fit of the predictive distribution to the data set (Figure 7). We observed that using the cumulative sum enabled the consideration of the uncertainty caused by uneven defecation frequencies. For example, the defecation frequencies of MO04 and MO12 were comparable, but the uncertainty was estimated to be larger for MO04 because of the uneven defecation frequencies.

**Figure 7:**
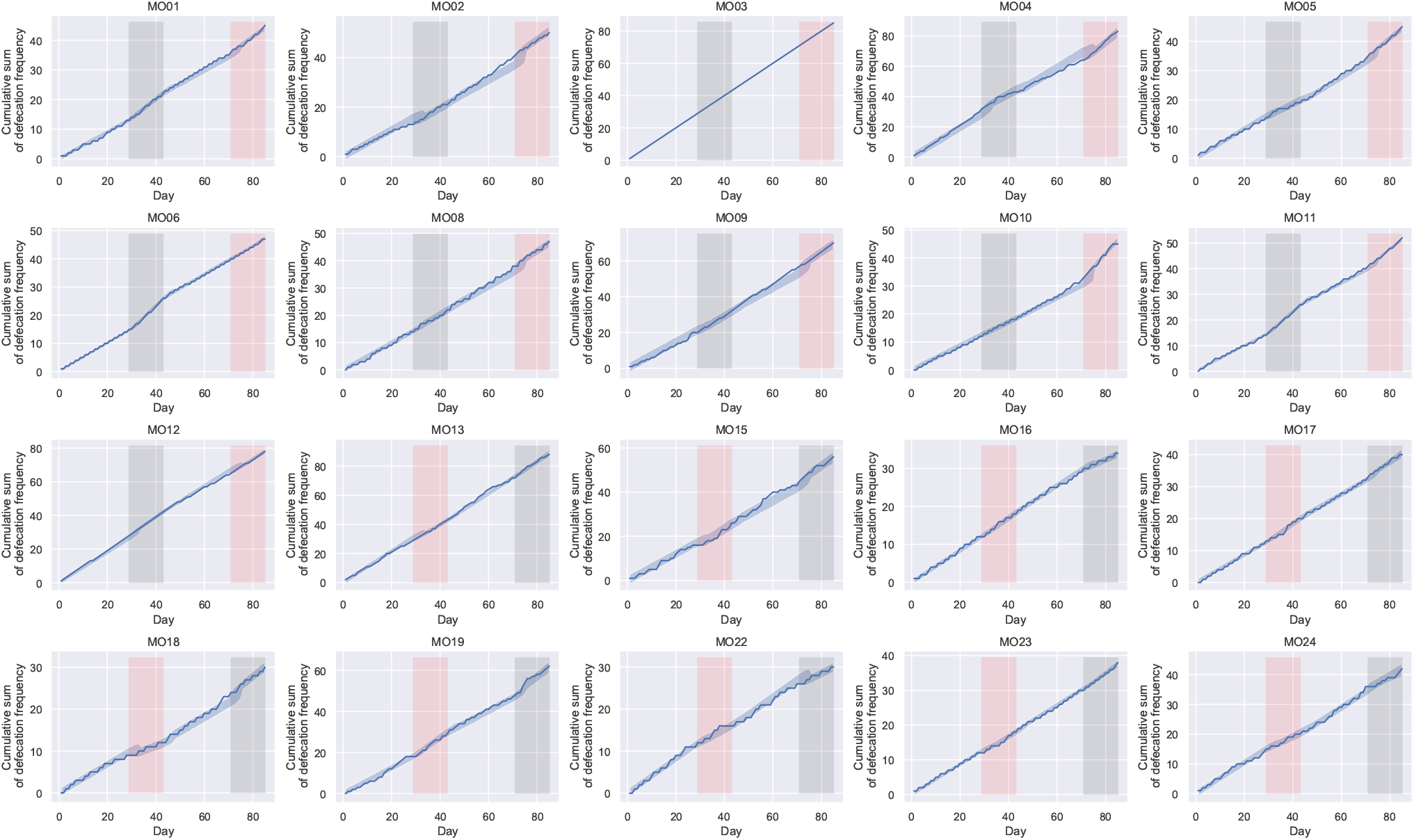
Data used for the analysis and predictive distributions. Each plot shows each subject. The *x*- and *y*-axes indicate the day and cumulative sum of the defecation frequencies, respectively. The blue line and the blue area indicate real data and the 2.5% and 97.5% percentiles of predictive distributions, respectively. The red and black areas indicate the target probiotic and placebo intake periods, respectively.

To investigate the relationship between the response to probiotics and gut microbiota, we performed Bayesian beta regression of the responder probability on the microbial abundance features before the target probiotic periods. Figure 8 shows the posterior distribution of the regression parameters. The negative effect of *Agathobacter* was estimated. This result for *Agathobacter* is consistent with that of the previous study [10]. However, The 95% Bayesian credible intervals for all regression parameters included zero, and we could not conclude any microbial effect based on them.

**Figure 8:**
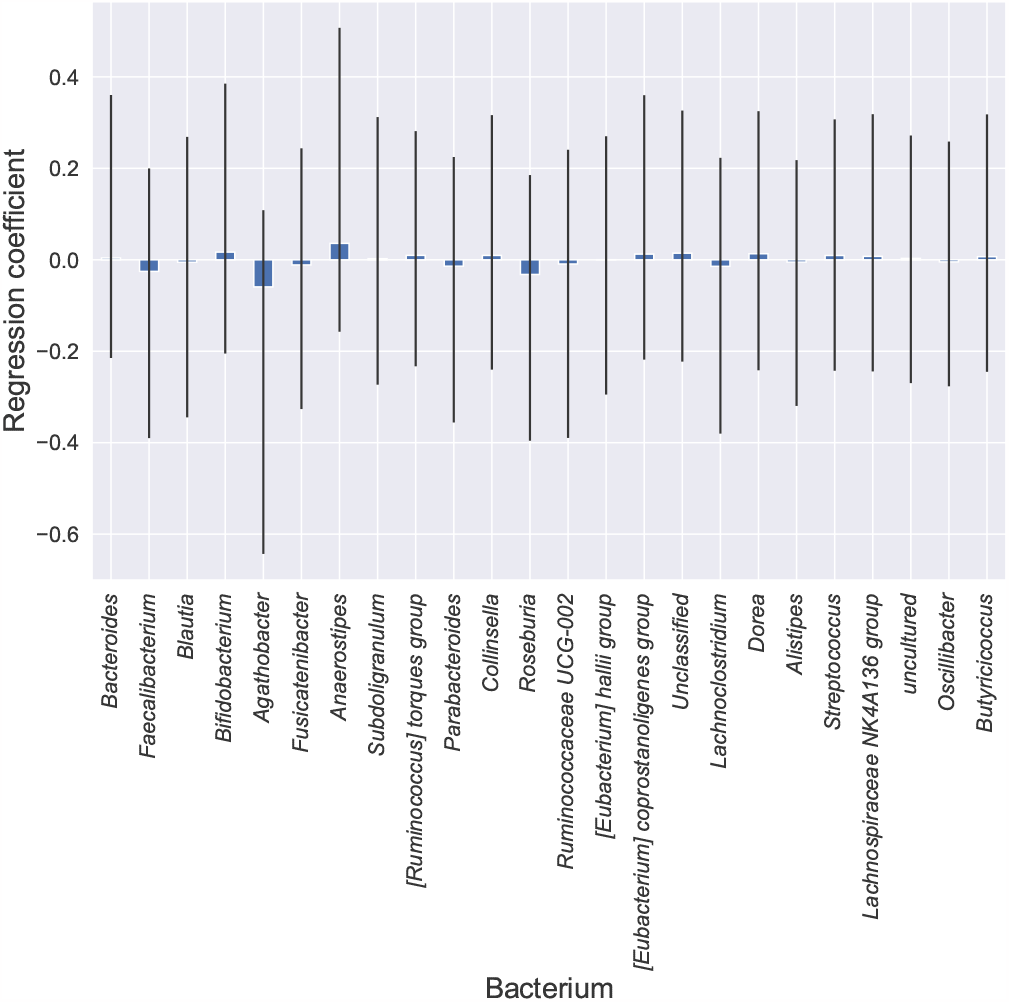
Posterior distributions of the coefficients of Bayesian beta regression. The *x*- and *y*-axes indicate bacteria and regression coefficients, respectively. The bar shows the median of the posterior distribution. The error bars represent the 2.5% and 97.5% percentiles.

## 4 Discussion

One of the benefits of using statistical Bayesian models is its adaptability and the potential for extensive customization. If our model is desired to be applied to other phenotype datasets, it can be employed with a slightly modified structure. For example, consider a blood pressure value case. *y*_*i*_ indicates the blood pressure value, which is continuous and not cumulative. In this case, the *y*_*i*_ distribution of the form

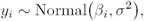

can represent the continuous values for days in each segment. Therefore, although the proposed model in this study was applied to the cumulative sum of defecation frequencies, the key idea of the model is supposed to be widely applicable in cross-over trial datasets.

We used the same lag of the start/end day for the placebo and the target probiotic periods. However, these two types of lags may differ because of their sources. While the lag in the target probiotic period is likely to be caused by physical factors (digestion and changes in physical conditions), the lag in the placebo period is likely to be caused by cognitive factors [21]. Therefore, introducing different lag parameters for the placebo and target probiotic periods may enable better estimation. However, adding these parameters can render the estimation computationally expensive.

We used uniform distributions with fixed hyperparameters as prior distributions of *μ* and *ν*. There are several options for the prior distribution. Setting prior distributions based on literature enables a more accurate estimation of parameters. In addition, the covariance between *μ* and *ν* can reflect the consistency of *μ* and *ν* in a subject if a covariate between *μ* and *ν* can be assumed.

*μ* and *ν* play key roles in the proposed model. In the synthetic data experiments (Section 3.1), the estimation performance of *μ* and *ν* was not very accurate. However, the proposed model is effective for estimating responders, as seen in the synthetic data experiments (Figure 3). This is because considering all cases of (*μ, ν*) contributes to the detection of responders when there is a lag in the effect period. However, *μ* and *ν* may not always be indispensable. The effects of *μ* and *ν* are limited for the long-term datasets because the number of lag days is small relative to the number of days in the trial. Indeed, the difference between the base model and the proposed model is smaller for synthetic long-term datasets containing observations made under similar conditions (Supplementary Figure S5 and Supplementary Figure S6). Nevertheless, consideration of lag is still significant due to the following reasons: first, the proposed model still outperforms the base model in long-term datasets (Supplementary Figure S5 and Supplementary Figure S6), and second, experiments are frequently limited to a brief duration for economic reasons.

There is a limitation to determining responders based only on defecation frequencies, which is suggested to be unreliable by the U.S. Department of Health and Human Services Food and Drug Administration [22]. According to them, the identification of responders needs to be evaluated based on defecation frequency and abdominal pain intensity. Therefore, deterministic estimation of responders may lead to wrong conclusions. We believe that the responder estimation based on the posterior distribution of *η* conducted in this study enables us to consider the uncertainty of the estimation and contribute to solving this problem.

## 5 Conclusion

In this study, we proposed a model for estimating the improvement in defecation frequencies using cross-over trial datasets and considering the lag between intake and effect periods. Using synthetic datasets, we verified that the proposed model could identify responders better than the base model. In the real dataset experiments, we identified seven responders based on the probability of *η >* 0. The base model, which assumed that the lag did not exist, identified subjects MO22 and MO24 as responders in addition to those identified by the proposed model. Subjects MO22 and MO24 did not have high DFI scores when *μ* ≠ 0 and *ν* ≠ 0 (Figure 6). The proposed model reflected these observations. The proposed model was suggested to eliminate uncertain responders regarding the lag between intake and effect periods. In the regression analysis of the responder probabilities on the microbial relative abundances before target probiotic intake, we found that *Agathobacter* had a negative effect. This result for *Agathobacter* is consistent with that of the previous study [10].

The proposed model effectively addresses the lag between intake and effect periods. The model demonstrated strong performance when lag was present. We believe that our model will help identify probiotic responders on cross-over trials for constipation.

## Supporting information

Supplementary data including Figures S1, S2, S3, S4, S5, and S6

## Author contributions

**Shion Hosoda:** Conceptualization, Methodology, Software, Investigation, Validation, Visualization, Writing-Original Draft. **Yuichiro Nishimoto:** Software, Investigation, Writing - Review & Draft. **Yohsuke Yamauchi:** Software, Investigation, Writing - Review & Draft. **Takuji Yamada:** Investigation, Writing - Review & Draft. **Michiaki Hamada:** Investigation, Supervision, Writing - Review & Draft.

## Acknowledgments

Computations were partially performed with the NIG supercomputer at ROIS National Institute of Genetics and the supercomputer at Human Genome Center, the Institute of Medical Science, the University of Tokyo. We would like to thank Editage (www.editage.jp) for English language editing.

## Data availability

Stan and Python source codes are available at https://github.com/shion-h/LagBasedResponderIdentifier.

## Supplementary data

Supplementary data, including Figures S1, S2, S3, S4, and S5, are available.

## Declarations

The randomized controlled trial whose dataset was used in this study was conducted with the approval of the clinical trial ethics review committee of Chiyoda Paramedical Care Clinic.

