## Supplementary data including Figures S1, S2, S3, S4, S5, and S6 for "Probiotic responder identification in cross-over trials for constipation using a Bayesian statistical model considering lags between intake and effect periods"

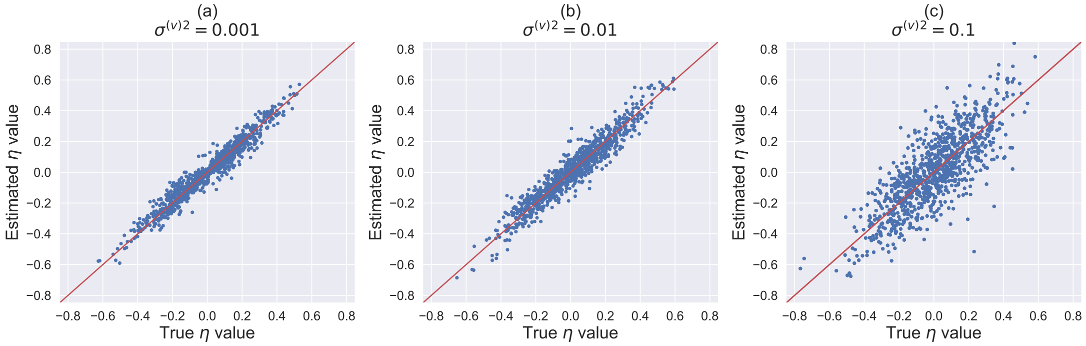


Figure S1: The scatter plots of the true $\eta$ and estimated posterior expectation of $\eta$ for each synthetic dataset, where the number of observation points was the same as that of the real dataset. The $x$- and $y$- axes indicate the true $\eta$ and estimated $\eta$ values, respectively. a, b, and c indicate ${\sigma^{(v)}}^2 = 0.001$, ${\sigma^{(v)}}^2 = 0.01$, and ${\sigma^{(v)}}^2 = 0.1$, respectively. The red line indicates that $y = x$.

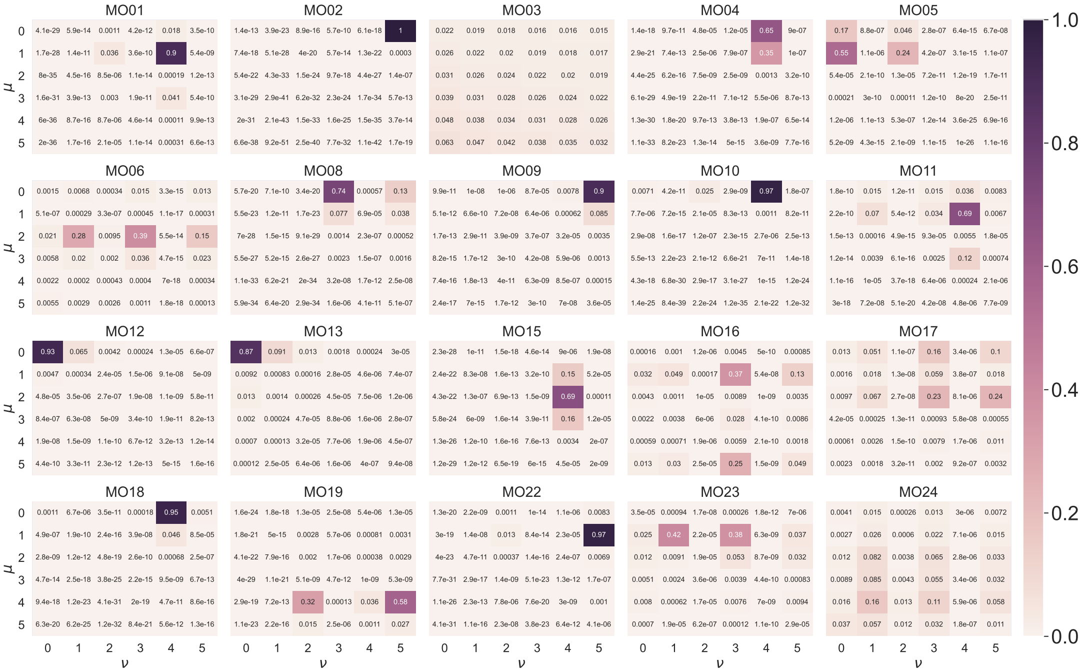


Figure S2: The heat map of the posterior distributions of $\mu, \nu$ for each subject. The $x$- and $y$-axes indicate the $\nu$ and $\mu$ values, respectively. All elements for one heat map sum to 1.

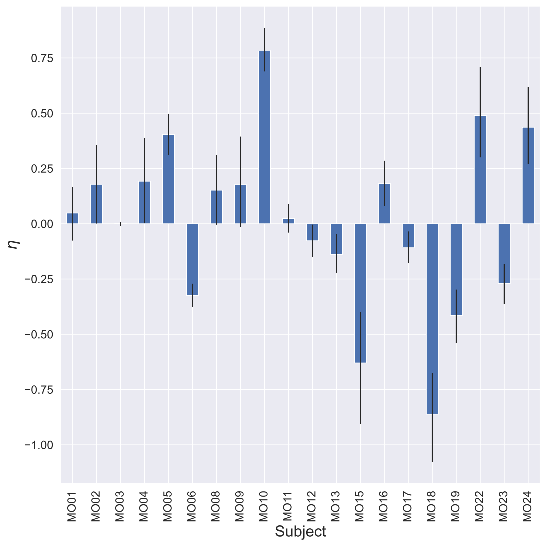


Figure S3: The posterior distributions estimated by the base model (*cf.* Section 3.1 in the main text) of $\eta$ for each subject. The legend is the same as in Figure 4 in the main text.

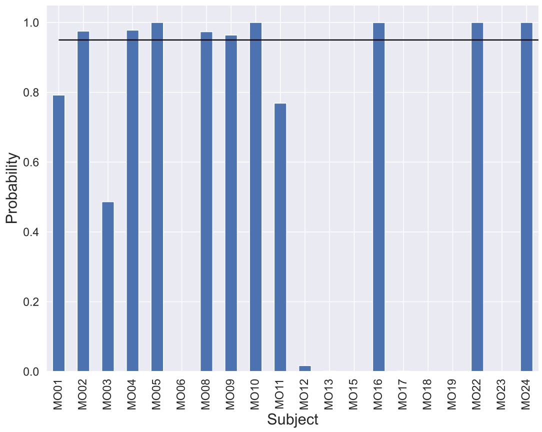


Figure S4: The probability that each subject is a responder based on the posterior distribution estimated by the base model (*cf.* Section 3.1 in the main text). The legend is the same as that in Fig. 5 in the text.

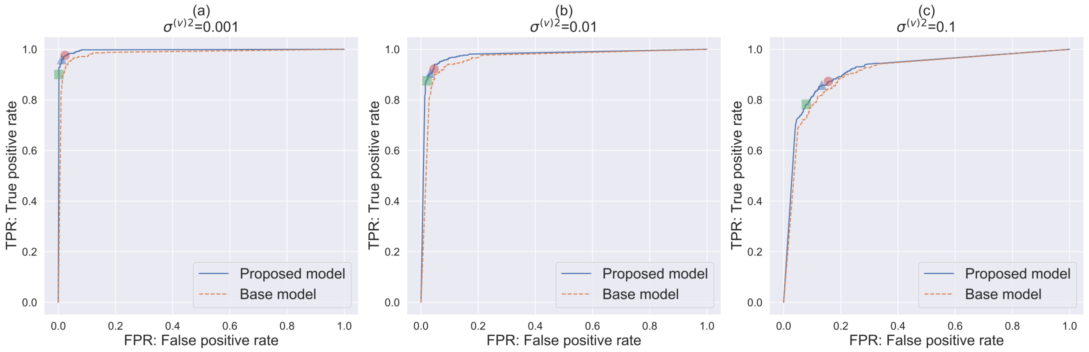


Figure S5: The ROC curve for identifying responders based on the estimated $\eta$ in the synthetic datasets of $N = 151$ and $(d_1, d_2, d_3, d_4) = (51, 76, 126, 151)$ using the proposed method when $\mu_{\max} = \nu_{\max} = 5$ and $\mu_{\max} = \nu_{\max} = 0$. The legend is the same as in Figure 3 in the main text.

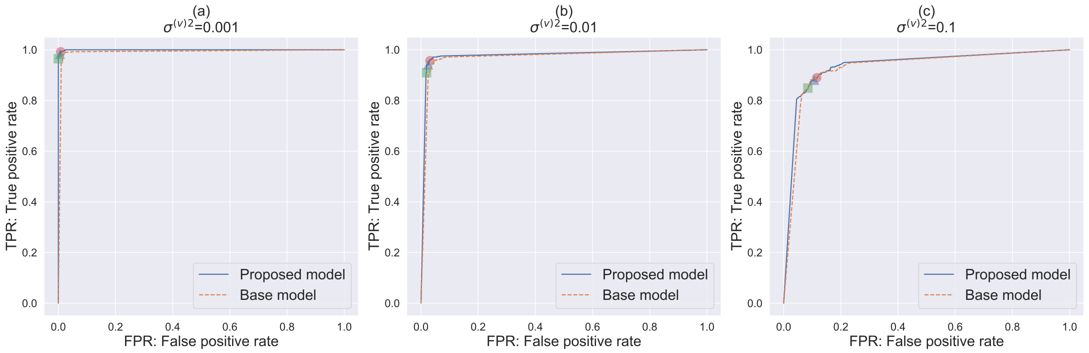


Figure S6: The ROC curve for identifying responders based on the estimated $\eta$ in the synthetic datasets of $N = 301$ and $(d_1, d_2, d_3, d_4) = (101, 151, 251, 301)$ using the proposed method when $\mu_{\max} = \nu_{\max} = 5$ and $\mu_{\max} = \nu_{\max} = 0$. The legend is the same as in Figure 3 in the main text.
